# Approaches to self-management integration and influencing factors in everyday life after spinal cord injury: A qualitative narrative analysis

**DOI:** 10.1101/2025.01.06.25320055

**Authors:** Enxhi Qama, Nicola Diviani, Clara Häfliger, Xavier Jordan, Anke Scheel-Sailer, Claudia Zanini, Sara Rubinelli

## Abstract

**Objective:** This study explores how individuals with spinal cord injury (SCI) integrate self-management (SM) into their everyday lives post-discharge from initial rehabilitation. It focuses on identifying the approaches they employ in balancing health tasks with personal and societal roles and the influencing factors.

**Methods:** We conducted semi-structured interviews with 32 participants, recruited from four rehabilitation centers across Switzerland, three months post-rehabilitation. Data collection spanned from November 2022 to May 2024. We used thematic analysis to identify the challenges and strategies associated with SM integration.

**Results:** Three distinct approaches to SM integration emerged: *The compartmentalizing approach,* where individuals focused on one aspect at a time; *The mixing approach,* where both health and other tasks were prioritized but adjusted; and *The embedding approach,* where there was equal prioritization with no adjustment on either side. Different factors such as mind and body dynamics, which encompass physical and emotional aspects; environmental and informational dynamics, which include external support, accessible facilities, and information; and society and perception dynamics, which include social stigma and misconceptions, influenced these approaches.

**Conclusions:** This study contributes to a more nuanced understanding of how to balance both medical and role management in SCI post-discharge. Self-management integration is achieved through different approaches and influenced by a wide range of factors, internal and external ones. Further research should longitudinally explore whether the approach one individual employs changes with the time and what aspects reinforce one or the other.

**Practical implications:** Our findings highlight the need for flexible, personalized SM interventions in SCI that are contextually grounded but also adaptive and resilient. Rehabilitation settings should assess different SM integration approaches, using feedback to guide individuals in refining their strategies. Communication guidelines and tailored education sessions are recommended to help align SM practices with patients’ evolving goals, including family, social, and leisure priorities.

## 1. Introduction

Spinal cord injury (SCI) affects between 250,000 and 500,000 people worldwide each year, impacting physical health, emotional well-being, social engagement, and personal relationships [1]. Effective self-management (SM) is essential for sustaining health and quality of life in complex chronic conditions like SCI [2, 3]. Self-management is defined as "the ability of the individual, in conjunction with family, community, and healthcare professionals, to manage symptoms, treatments, lifestyle changes, and psychosocial, cultural, and spiritual consequences of health conditions" [4].

Self-management interventions for SCI typically address medical practices such as catheterization, bowel care [5, 6], skin checks [7, 8], mobility, physical activity [9, 10], and medication management [11, 12]. These interventions have demonstrated benefits in reducing pressure ulcers [9], urinary infections [10], as well as improving self-efficacy [11] and functional independence [12].

Despite these benefits, two key limitations persist. *First*, most SM programs are implemented in community settings with individuals already living with SCI for years [13], with fewer interventions targeting the critical transition from rehabilitation to community life [14]. This phase greatly impacts the long-term adjustment process following the acquisition of SCI [15-17], characterized by high prevalence of secondary complications and rehospitalization [18-23]. *Second,* many SM interventions focus primarily on the medical aspects, overlooking the evidence that SM is far more complex and extends beyond symptom management [24, 25]. Successful SM requires balancing diverse health tasks alongside evolving life circumstances [26-28], such as family responsibilities, social participation, and employment [6, 7, 29-33]. When this balance results in failure, evidence shows that it can lead to two major challenges: poor adherence and low sustainability of health management over time [34-37], and dissatisfaction and low participation in leisure activities, domestic life, and relationships, even years after community reintegration [31, 32, 38]. Therefore, it is particularly important to understand the transition period from rehabilitation to everyday life [14], the role it plays in building the skills people need to start managing their disability [20-22, 39], and how they apply and adapt the SM information they receive [40-42]. Consider the management of pressure relief [43] or the execution of catheterization procedures during travel [28].

To address these gaps, this study aims to explore how individuals with SCI integrate SM into their lives after discharge from initial rehabilitation. Specifically, it focuses on the approaches to SM integration they follow and the influencing factors. Understanding these aspects could inform the development of targeted SM interventions that consider the lived experiences of individuals. Ultimately, this will help improve the long-term sustainability of SM for those affected by SCI.

## 2. Methodology

### 2.1 Study Design

We chose a qualitative approach in order to have a deep understanding of how individuals experience the phenomena of SM [44, 45]. Specifically, we applied an experiential orientation since we want to understand the lived experiences and personal meaning-making of SM [46]. This study addresses two research questions:

**RQ1:** What approaches do individuals follow in integrating SM practices into their everyday lives following discharge from initial rehabilitation?

**RQ2:** What factors influence the SM integration?

We used the Standards of Reporting Qualitative Research (SRQR) checklist to report our study [47] (see Supplementary file 1).

### 2.2 Setting and participants

This study was conducted within the Swiss Spinal Cord Injury study (SwiSCI), a multi-center, longitudinal inception cohort study. SwiSCI aims to enhance understanding of how to support functioning, health maintenance, and quality of life for individuals with SCI across the continuum of care, from rehabilitation to community life. The study includes individuals aged 16 or older residing in Switzerland with traumatic or non-traumatic SCI, excluding those with congenital conditions (e.g., spina bifida), neurodegenerative disorders, or new SCI in palliative care. SwiSCI is structured into three pathways, with this study embedded in Pathway 3, which follows individuals with newly acquired SCI. Detailed information on SwiSCI is available elsewhere [48].

We recruited participants from four rehabilitation centers in Switzerland who had completed their rehabilitation program and were under the care of an interprofessional rehabilitation team, which provides training and instructions. We interviewed individuals recently diagnosed with a SCI three months after discharge.

A Swiss research assistant of the SwiSCI study informed potential participants about the study at the time of discharge and gave them a flyer as a reminder. The participants (n=50) who expressed interest provided their contact details (email address or phone number), and within a month, a researcher (CH) contacted them via email or phone to provide a detailed explanation of the study, including their rights and obligations. We stopped further outreach if the first three contact attempts proved unsuccessful. We scheduled interviews three months post-discharge and obtained informed consent from participants (n=34) who maintained interest (see Figure 1 for the recruitment flowchart).

**Figure 1.**
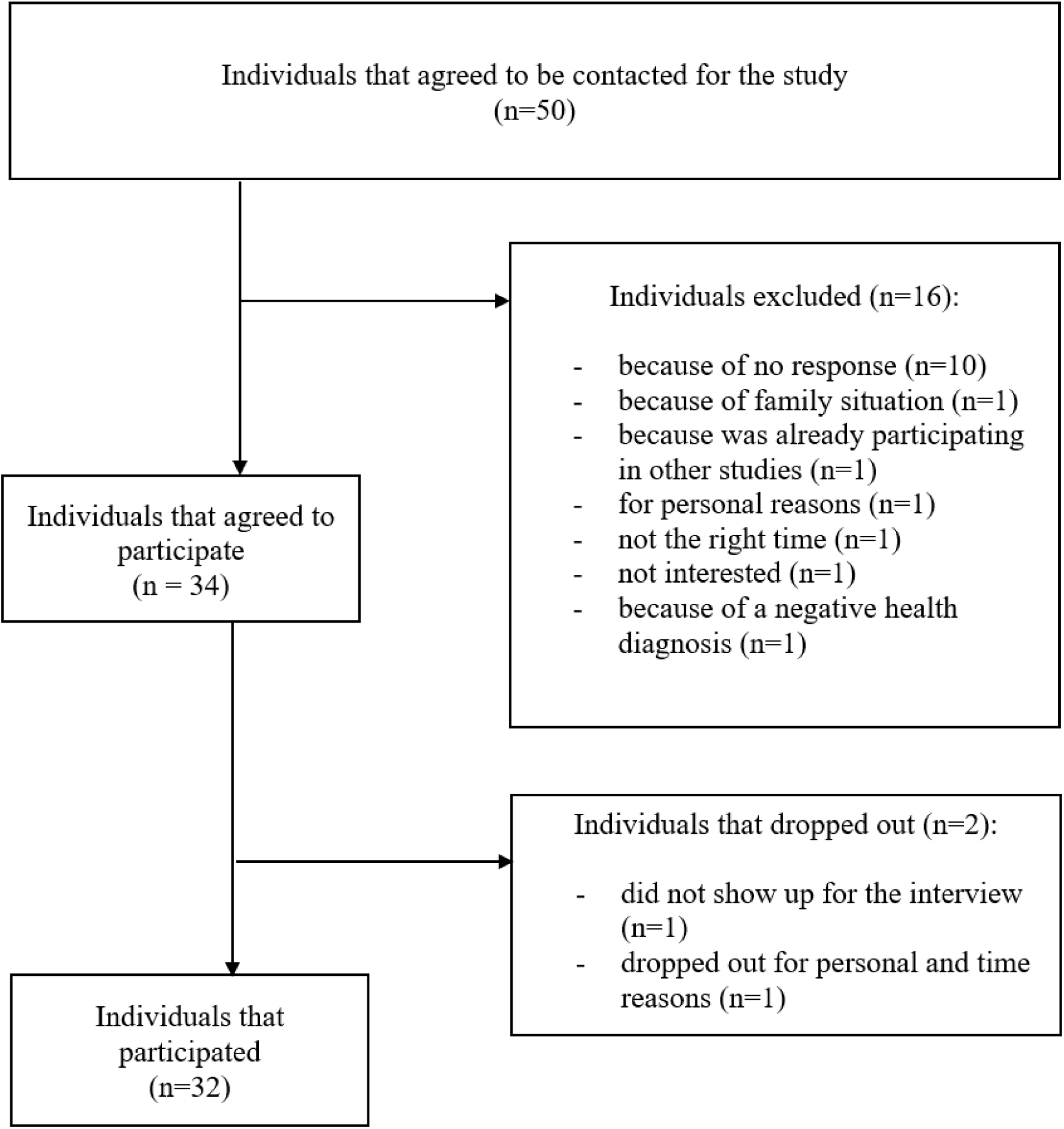
Recruitment flowchart.

We continued with recruitment until there was the most variety in the types of injuries (complete/incomplete) and levels of paraplegia/tetraplegia, as well as thematic saturation from the last five interviews that were analyzed (see Table 1 for details on the participants).

**Table 1.** Socio-demographic of participants.

| **Characteristic (N=32)** | **n (%)** |
| --- | --- |
| **Age** |  |
| Range | 19-78 |
| Median | 49 |
| **Sex** |  |
| Female | 4 (12.5%) |
| Male | 28 (87.5%) |
| **SCI level** |  |
| Paraplegia | 17 (53%) |
| Tetraplegia | 15 (47%) |
| **Severity of injury** |  |
| Complete | 8 (25%) |
| Incomplete | 24 (75%) |

### 2.3 Data Collection

We conducted semi-structured interviews from November 2022 to May 2024. Our interview guide broadly covered various facets of SM by incorporating our understandings from the COM-B behavior model [49-51]. This model, which explains how capability, opportunity, and motivation shape behavior [52], is particularly significant in the context of SM. Successful SM, such as managing bladder or bowel issues for individuals with SCI, requires a synergy of capability (knowledge and skills), opportunity (access to health services and support), and motivation (will and commitment) from the individuals’ side [53, 54].

The interview guide also drew from previous research that explored diverse settings and situations in daily life where SM of complex chronic conditions is contextualized and naturalized. This includes areas such as work, leisure activities, and social participation [29, 55]. A research professional (CH, CZ) with experience and training in qualitative interviews conducted the interviews in the native language of the interviewees and pilot tested the guide.

We started the interviews by asking participants about their general thoughts on SM activities related to their health situations. We then explored their experiences during and after initial rehabilitation. Finally, we asked the participants to reflect on how they applied their acquired knowledge in their daily environments. We used probes to elicit deeper responses (refer to Supplementary file 2, for the interview grid). We conducted interviews in person (at the participant’s chosen location), via phone, or Zoom [56, 57], with durations ranging from 21 to 82 minutes. The research team only had access to an internal folder containing audio recordings of all interviews. We took field notes during each session (CH, CZ). A researcher (EQ) also wrote reflective summaries to aid in the data analysis.

### 2.4 Data Analysis

We employed thematic analysis, which involved a deep reflection on the data and engagement with participants’ stories [58, 59]. The process began with familiarization, including repeated listening to the recordings and transcript readings. We cross-referenced the initial notes with the preliminary impressions formed during the readings. One researcher (EQ) conducted the initial coding using MAXQDA software, applying broad and descriptive codes (open coding) [59]. Next, EQ and ND collaborated to enhance and arrange the codes through axial coding, emphasizing the methods and elements of SM integration (refer to Figure 2 for an illustration of the development of codes and themes).

**Figure 2.**
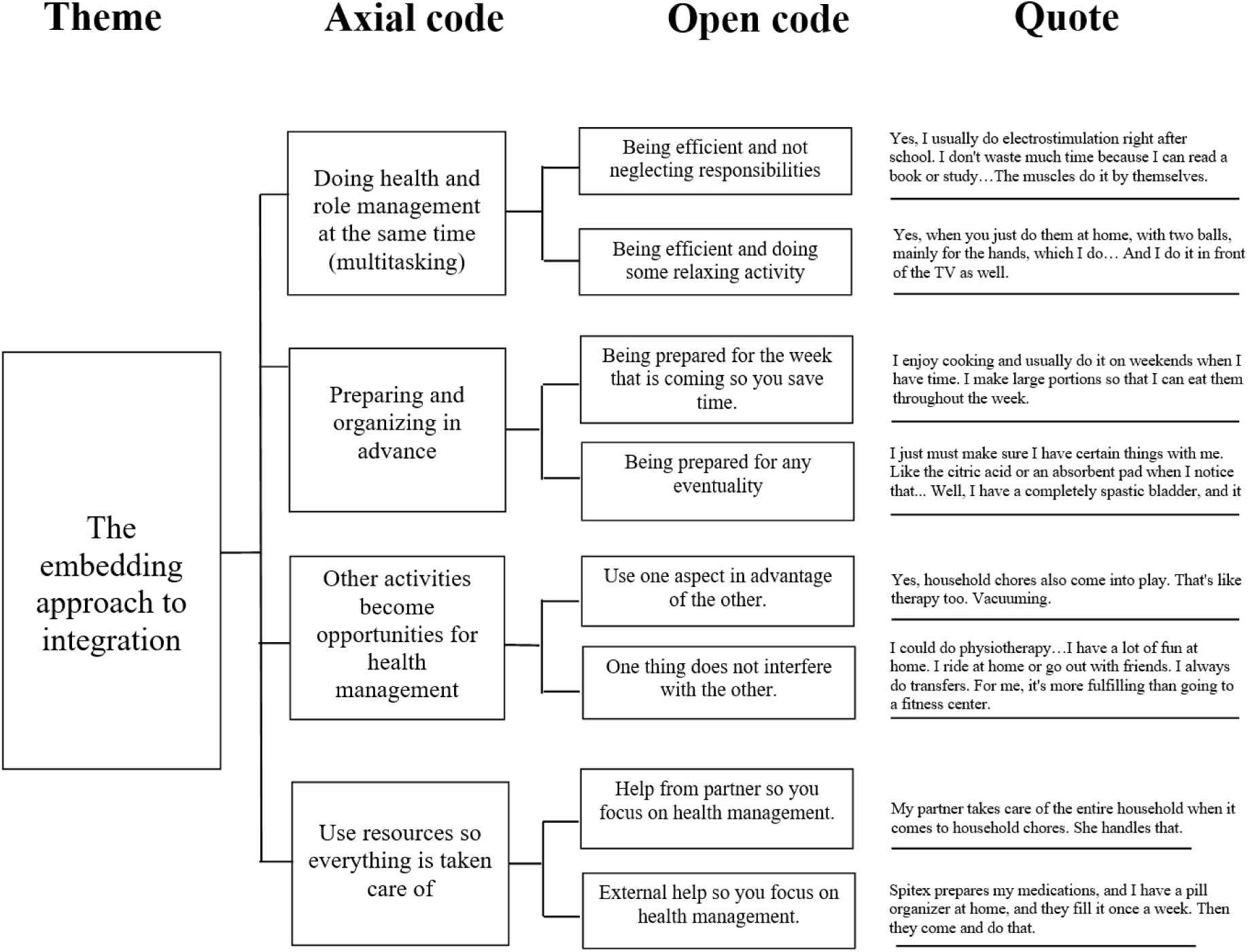
Illustration of code and theme development.

We defined approaches as ways individuals experience SM integration, influenced by their circumstances, capacities, and readiness. We defined factors influencing SM integration as elements that either hinder or facilitate individuals’ ability to integrate SM practices into their everyday lives.

In the final stage, four researchers (EQ, ND, CH and CZ) collaborated on theme generation that reflected a process of intertwining the data themselves and the researchers’ analytical expertise seeking consensus about meaning, aiming for a nuanced understanding of the data [45, 58, 60]. To enhance trustworthiness, we employed strategies like field notes and reflective summaries [61]. Additionally, the interviewer was a native speaker and not involved in the healthcare of the participants, fostering an environment of trust and open dialogue. The research team was experienced in qualitative research and had some of the members had health backgrounds as a speech therapist and pharmacist. Trustworthiness of the analysis was further ensured through discussions at university seminars and international conferences [62].

### 2.5 Ethical Considerations

The study obtained ethical approval from the regional committee (ref. EKNZ 2022-00501). Additionally, SwiSCI adheres to national and international research standards, including the Declaration of Helsinki by the World Medical Association (World Medical Association 2008), the "International Ethical Guidelines for Epidemiological Studies 2009" by the Council for International Organizations of Medical Sciences (Council for International Organizations of Medical Sciences 2009), and national guidelines for research integrity (Akademien der Wissenschaften Schweiz 2013).

## 3. Results

### 3.1 Approaches to SM integration

This study identified three distinct themes as approaches to SM integration: *The compartmentalizing approach*, *The mixing approach,* and *The embedding approach*. We provide a detailed description below, and Supplementary file 3 offers an extensive list of quotes for each approach.

#### 3.1.1 The compartmentalizing approach to SM integration

This approach reflects participants’ commitment to addressing either health-related SM tasks or other life activities as separate priorities, depending on the situation. For instance, some participants view bowel and bladder management as essential to prevent complications, which motivates them to dedicate significant time to hygiene practices and prioritize health over social or leisure activities (Table 1, Q (1-2)). The same applies for physical recovery (Table 1, Q (3-4)). However, personal responsibilities, such as parenting, sometimes necessitate a delay or set aside of SM activities to attend to family needs (Table 1, Q5). Some participants feel that social outings or vacations justified a shift in focus, canceling therapies to engage in these events fully (Table 1, Q (6-7)).

#### 3.1.2 The mixing approach to SM integration

The mixing approach is characterized by a flexible and adaptive attitude toward SM integration. Here, the focus is on both SM tasks and other life activities, with participants adjusting them based on the circumstances, available time, and type of activity. They often modify the frequency to fit their daily lives, such as by experimenting with the timing of catheterization (e.g., starting at 6 a.m. or 2 a.m.) or adjusting exercise routines (e.g., daily or twice a week) to meet their physical needs (Table 2, Q (1-2)). Participants also adapt techniques to make SM tasks more manageable such as simplifying decubitus prevention steps, depending on the home environment or adjusting food preparation (e.g., preparing simpler meals) to save time and reduce effort (Table 2, Q (3-4)). Schedules are often rearranged to balance SM with other aspects of life, postponing therapy sessions or delaying pressure relief when social engagements or other activities take priority (Table 2, Q (5-7)). In some cases, participants could even decide to skip SM activities entirely for rest or personal time, recognizing the need to take occasional breaks from the heavy demands of therapy (Q9).

#### 3.1.3 The embedding approach to SM integration

Participants in this SM integration approach maintain both health tasks and daily life activities without adjusting or compromising on either. Multitasking is a common strategy here, such as studying while doing electrostimulation, watching TV while exercising, or catheterizing during car journeys to avoid restroom stops (Table 3, Q (1-3)). Preparation also helps participants to maintain control over their health without having to sacrifice their regular routines. They organize tasks to avoid disrupting their daily routines, such as using medication organizers, batch-cooking meals for the week, and carrying items like pads or urine bags for outings (Table 3, Q (4-8)). Participants also use enjoyable or essential daily activities, like hobbies or household chores, as opportunities to keep up with SM exercises (e.g., engaging in a hobby that involves movement or using chores as physical activity) (Table 3, Q (10-12)). Finally, when tasks are too challenging or time-consuming, participants rely on family or professional help to ensure SM and daily responsibilities are managed (e.g., having partners prepare meals or support services organize medications (Table 3, Q (13-15)).

### 3.2 Factors influencing SM integration

This study identified three themes regarding factors influencing SM integration: *Mind and body dynamics*, *Environmental and informational dynamics,* and *Society and perception dynamics.* We provide a detailed description below, and Supplementary file 4 offers an extensive list of quotes for each factor.

#### 3.2.1 Mind and body dynamics influence SM integration

The physical and emotional demands shape how individuals approach SM integration, influencing both their capacity and strategy for managing health. Physical demands, particularly energy management, affect SM integration for individuals with SCI. Medication side effects and daily activities often diminish energy reserves, requiring participants to carefully plan their day to avoid complete exhaustion. This may involve adjusting work hours and scheduling downtime to maintain functionality for later tasks (Table 1, Q1).

Emotional and cognitive factors also play a key role in integrating SM into daily routines. Participants noted the substantial "brain power" needed to manage new routines, especially when adapting to major life changes. This mental load impacted their ability to stay focused on SM tasks, resulting in lapses in practice. Additionally, psychological challenges, such as motivation and navigating personal limits, added complexity to decision-making around SM, requiring individuals to continuously assess and balance what was feasible and desirable within their day-to-day lives (Table 1, Q (2-3)).

#### 3.2.2 Environmental and informational dynamics influence SM integration

This group of factors encompasses external elements that play a role in shaping SM integration. Practical aspects, such as living arrangements and support availability, impact the ease of managing health tasks. For instance, living alone often necessitates handling all tasks independently, adding to the complexity of SM routines (e.g., household chores and managing health apart). Conversely, some participants benefit from specific home modifications that facilitate mobility and routine tasks, such as wider showers and accessible kitchens. However, reliance on home care services sometimes introduces challenges, as services often operate on fixed schedules that restrict personal freedom and spontaneity, disrupting participants’ routines such as waiting for home care visits in the evenings (Table 2, Q (1-8)).

Informational and social support also proved essential in SM integration. Many participants find value in peer networks and ongoing connections with rehabilitation groups, offering both emotional support and practical advice on managing SCI and day-to-day challenges (e.g., sharing tips in WhatsApp groups, or keeping up with peer discussions even after rehabilitations). Additionally, some expressed the need for guidance and information on topics often overlooked in medical settings, such as managing body temperature during seasonal changes (e.g., sweating in summertime) and maintaining intimate relationships (e.g., getting the partner to attend workshops). (Table 2, Q (9-12)).

#### 3.2.3 Society and perception dynamics influence SM integration

This group of factors encompasses societal perceptions and misconceptions around SCI, shaping how individuals approach and experience SM integration. In professional settings, accommodating SCI-specific needs (e.g., extended restroom breaks or rest periods) is sometimes met with understanding but can still underscore the challenges of balancing health management with work responsibilities. Although some workplaces offer flexibility, individuals may feel the need to manage perceptions around their unique needs, affecting their comfort and confidence in these settings (Table 3, Q (1-2)).

Social interactions around SCI also reveal gaps in societal understanding. Individuals are questioned about their need for mobility aids if they show any physical ability (e.g., going from a wheelchair to crutches), or they may feel hesitant to explain the realities of managing SCI needs like catheterization (e.g., friends or colleagues showing reluctance). Such interactions can create barriers to open social engagement, making individuals feel as though they must constantly justify their routines or downplay certain SM practices to fit societal expectations (Table 3, Q (3-6)).

## 4. Discussion

This study aimed to explore the personal experiences of SM integration in individuals with SCI by uncovering both their approaches and influencing factors.

**Regarding the approaches,** the analysis revealed three different ways of SM integration, with three distinct focuses. In the compartmentalizing approach to integration, individuals concentrate <u>on one priority at a time</u>; in the mixing approach, they focus on <u>both priorities with modifications</u>; and in the embedding approach, they concentrate on <u>both priorities without any adjustments</u>. Research in SCI and other chronic conditions [63] has similarly explored the balancing act between managing a chronic disease and living a meaningful life [24-27]. Zanini et al., for instance, describe the flexibility in prevention of pressure ulcers, showing the different styles that people apply, sometimes by delegating and sometimes by being selective with their priorities [8]. Our study, however, provides novel insights by examining SM as a multifaceted process that goes beyond isolated medical tasks. We emphasize SM as the integration of health management as well as personal (e.g., parental roles, household tasks), social (e.g., vacations, outings), and professional responsibilities.

**Regarding the influencing factors**, we identified distinct groups ranging from personal to external (e.g. resources and social perceptions), similar with previous research [64-67]. Our study extends these findings by show that understanding one’s body is crucial not just for specific health-related tasks but for all aspects of daily life activities. For instance, bodily limitations or energy levels, such as fatigue or emotional fluctuations, can interfere with the decision of choosing health or daily activities. Informational resources are crucial, particularly when it comes to intimacy and sexuality, yet existing SCI interventions often overlook this SM aspect [68]. Lastly, the stigma associated with invisible disabilities, such as incomplete SCI, or SM tasks like bladder and bowel care, creates barriers for SM integration, which often go unnoticed in intervention design [13].

Our study has some important strengths. *First*, it presents a novel perspective on SM, extending beyond isolated medical tasks [6] to explore how SM is embedded in daily life [63], balancing personal, social, and professional responsibilities. *Second,* this is the first study, to our knowledge, to explore SCI patients’ experiences with SM integration post-discharge from initial rehabilitation. *Third,* the qualitative approach allowed for an in-depth exploration of individuals’ lived experience, offering rich, nuanced insights. Our use of maximum variation sampling ensured that the findings reflect a diverse range of experiences across different injury levels and demographics, enhancing the transferability of the results. *Finally,* our study adhered to the Standards of Reporting Qualitative Research (SRQR), ensuring transparency and replicability. By thoroughly documenting our data collection and analysis procedures, we offer a robust and reliable framework that can guide future research in this area.

Several limitations must be acknowledged. *First,* generalizability may be limited as this study was conducted with participants living in Switzerland, where healthcare policies, insurance structures, and rehabilitation services [69] may differ from other countries [70]. However, we included participants from four rehabilitation centers across Switzerland, which already provided some diversity in terms of cultural and environmental factors, even within the same country. *Second,* the language restriction in our recruitment criteria may have excluded non-Swiss (German, French, or Italian) or English speakers. This means that we may have missed valuable perspectives from non-Swiss speakers, such as immigrants and expatriates, who could have contributed valuable perspectives on SM integration, particularly regarding language barriers and navigating a foreign healthcare system. *Third,* we do not have knowledge of the health behaviors and lifestyles the participants had before their SCI and whether they influenced their approach to SM integration post-injury.

Our study sheds light on several areas for further exploration. Firstly, we cannot definitively assign the identified approaches to any individual. People’s approaches may vary depending on the nature of the activity they’re discussing (e.g., whether it’s a delegable or time-critical task) or their individual circumstances, abilities, or readiness [71]. Even though the embedding approach seems to be the ideal approach, we need longitudinal research to track if there is an evolution of SM integration over time, if individuals shift between approaches, and also what they would need to develop the most appropriate approach for their circumstances. Second, further research on problem-solving and decision-making skills [11, 72]—particularly as individuals encounter evolving circumstances and external factors— could provide valuable insights into the mechanisms that fuel SM integration [73]. Additionally, research should investigate whether different subgroups of individuals with SCI experience SM integration differently (e.g., by injury level, social background, or cultural factors). This could uncover the likelihood of certain groups encountering specific approaches and guide the development of tailored interventions to cater to the diverse needs of individuals with SCI.

## 5. Conclusion

There are different approaches applied for SM integration, as well as diverse influencing factors. The insights of this study contribute to a more nuanced understanding of how to balance both medical and role management in SCI post-discharge. Supporting individuals in this process requires interventions that are contextually grounded but also adaptive and resilient—continuously evaluated and appraised over time. Additionally, this study contributes valuable insights for future research, particularly by identifying the need to longitudinally track changes in an individual’s SM journey after discharge from rehabilitation.

### 5.1 Practice implications

By emphasizing the importance of adaptation and situating health tasks within the broader context of daily life, our findings call for more flexible and personalized SM interventions that go beyond traditional informational modules [74]. Adherence to these interventions is not solely a matter of following instructions but rather of accommodating the diverse and dynamic realities individuals face [75-77].

Rehabilitation settings, which are critical for long-term adjustment to SCI [78-80], should focus on assessing the specific approaches to SM integration that individuals are taking. Monitoring their outcomes can provide valuable feedback, enabling health professionals to help individuals appraise and, if necessary, re-evaluate their approaches. Depending on the rehabilitation and support programs in place, health professionals or peers may administer follow-up visits, evaluation interviews, or dedicated assessment tools to facilitate this continuous process of appraisal [81]. This personalized evaluation would take into account the influencing factors relevant to the individual’s circumstances at a given point in time.

Effective communication between health professionals and patients becomes essential in promoting successful SM integration [82, 83]. Health professionals must communicate treatment plans [77, 81] but also offer the right guidance and discharge education in order for SM tasks to realistically fit into daily life. For example, health professionals should ensure that SM aligns with what is most meaningful to the patient, which often extends beyond functional independence to include participation in education, family responsibilities, and leisure activities [84, 85]. It’s also important to recognize that patients’ priorities may shift over time, particularly within the first years after injury [55]. Communication guidelines highlighting best practices for discussing SM approaches, along with education sessions focusing on SM integration, could achieve this by making health tasks more approachable for individuals.

## Data Availability

All data produced in the present work are contained in the manuscript

## Funding sources

This work was supported by the Swiss National Science Foundation (www.snf.ch; Grant No. 10001C_200520). The funding source had no role in the study’s conceptualization, decision to publish, or preparation of the manuscript.

## Author statement

Conceptualization: Enxhi Qama, Nicola Diviani, Sara Rubinelli. Data curation: Enxhi Qama, Clara Häfliger. Formal analysis: Enxhi Qama, Nicola Diviani, Clara Häfliger, Claudia Zanini. Funding acquisition: Nicola Diviani, Sara Rubinelli. Investigation: Enxhi Qama, Clara Häfliger, Claudia Zanini. Methodology: Enxhi Qama, Nicola Diviani, Sara Rubinelli. Project administration: Enxhi Qama, Nicola Diviani, Clara Häfliger. Supervision: Nicola Diviani, Sara Rubinelli. Validation: Nicola Diviani, Sara Rubinelli. Writing – original draft: Enxhi Qama. Writing – review and editing: Enxhi Qama, with input from all co-authors.

## Notes

### Competing Interest Statement

The authors have declared no competing interest.

