## Supplemental_2 for "Approaches to self-management integration and influencing factors in everyday life after spinal cord injury: A qualitative narrative analysis"

**Table 1. Sample questions for the semi-structured interview**

| Topic | Sample questions |
|---|---|
| **Introduction and warm-up** | • Are there any activities for self-management and prevention of complications of your SCI that you perform regularly? If yes – can you name them?<br>• Do you feel that you are keeping up well with these requirements? Why (not)?<br>• How much would you say does SM interfere with your life? Why? How? |
| **Initial rehabilitation** | Thinking back to the time you were in first rehabilitation and more specifically, to when you were taught about SM activities.<br>• How was it for you learning the new behaviors of taking care of yourself?<br>• What was particularly useful and what, instead, was not?<br>• Is there anything you think you should have been taught in first rehab but was missing? |
| **Return to the community, and integration of SM** | Now think back to when you first went back to the community (home, etc.)<br>• How was the implementation at home of the routines and SM regimes?<br>• Can you describe a situation regarding SM that was difficult for you to manage? Why? What made it difficult for you to?<br>• Have you made any compromises so far, because of SM activities you have to perform? Were there any situations when you had to decide either to engage in SM or to do other activities?<br>• Did you find a way to integrate these SM activities to your lifestyle? What did you do to take control? How? What did you need? |
| **Personal considerations** | • Is there anything else that is particular to your situation that helps you to better self-manage your condition? |

SCI – Spinal cord injury
SM – Self-management
