## Supplemental_3 for "Approaches to self-management integration and influencing factors in everyday life after spinal cord injury: A qualitative narrative analysis"

**Quotes representing three different integration approaches**

**Table 1. The compartmentalizing approach**

| **Q (1-2)** |
|---|
| "The essential thing is bowel management, emptying, etc…that's very uncertain. So, I took the risk. I would take the risk that I would be dealing with bowel management all day, and nothing else. That's of course bad… And I notice that this is something that affects my mood the most." (P26)<br><br>"Of course, there are many things I used to enjoy doing. But that's not a topic for me anymore…You can easily imagine that. When you watch motocross, in [name of a city], for example, there comes a point where you have to catheterize or use the portable toilets…That's why it was more important for me to maintain good hygiene… then risking a potential bladder infection." (P14) |
| **Q (2-4)** |
| "But the focus was on rebuilding physically. The other aspect is secondary" (P15)<br><br>"That's right. My health is my top priority. I didn't find it difficult to give up such plans. We have friends who come over, but I never go out. How could I? It's impossible…" (B11) |
| **Q (5-7)** |
| "…at 6 p.m. [at rehabilitation] it's time to relax. I could lie down, watch TV. That didn't work at home. I can't tell my little one, "I'm going now." (P6)<br><br>"I've canceled MTT twice because I wanted to do something social. Which gives a different kind of pressure, it's not just one thing, it's also the social aspect. And most people can do it in the evening, not during the day. Prioritizing MTT, but if it doesn't work out otherwise, then it gets shortened." (P30)<br><br>"I: Did you have to cancel any therapies during that time? Or were there no therapies scheduled? P: No, I canceled them for a week. It was only for a week…Mr. [name of doctor] said that during that week, you will learn more than we could teach you in a few weeks" (P10) |

**Table 2. The mixing approach**

| **Q (1-2)** |
|---|
| "I started catheterizing at 6 a.m. and 2 a.m... You have to experiment. You have to move things around... If it works for you, you can do it that way." (P16)<br><br>"They showed me enough exercises, in great detail. Ultimately, it's up to the individual how much you actually do them at home. If you do them daily or just twice a week, I sometimes catch myself thinking, "I've done enough today, and I don't feel like lying on the mat for another half hour doing exercises," (P19) |
| **Q (3-4)** |
| "So, they gave me a basic package or a basic set of instructions that covered everything I |

| |
|---|
| needed to know. And then I tried it out. At first, I tried it out exactly as they said. But I thought that it wouldn't work for me at home the same way. So, I tried to figure out how I could make it easier for myself. So, I think they did well in that aspect. I got all the information I needed there. They also mentioned multiple times and quite intensively that decubitus prevention was something I really had to pay attention to." (P7)<br><br>"Yes, the main thing that has changed significantly for me lately, especially compared to before, is cooking… I'm trying to keep things simpler and make meals that are less elaborate or require less chopping. So, I aim for something that's not too time-consuming or complicated, rather than just stirring things in a pan." (P19) |
| **Q (5-7)** |
| "Occasionally doing something with colleagues. And there, I feel like I have to decide what to do…for example, MTT…The advantage is that I don't have fixed MTT times…That's a big advantage. Then you can easily reschedule without any problems." (P6)<br><br>"Yes, it can be stressful. If I realize I've been in my wheelchair for two hours without shifting, because I'm out with people and haven't relieved pressure yet, I should have relieved pressure by now. I should have relieved it when I had the chance. But sometimes, when you're out and about, you don't have the opportunity, even though you know you should. It's like, "I can't do it right now." But if it's possible when I'm home, I'll tell myself to lie down for a moment, to shift my position. It's essential because later, if I want to go for lunch and then have physiotherapy, I won't have the time." (P7)<br><br>"I: Were there situations where you decided against self-management?<br>P: No, not really. That I delayed it, that happened. We learned to catheterize every four hours and at 7:00 the first time. Over time, I postponed it more and more. If I wanted to sleep longer, I did bowel management in the afternoon." (P21) |
| **Q (8)** |
| "You have to find a daily routine and it can't just be 100% therapy, otherwise you might as well stay in the rehab facility…I would say the biggest risk I've seen so far is perhaps the balancing act between how much I do, because, you know, the physiotherapist also says that I need to push these muscles a bit to the maximum so that they get triggered and make progress. Physiologically, I'm aware of that. But at the same time, being able to say, you know, no, now I'll take half a day off." (P13) |

MTT-medical training therapy

**Table 3. The embedding approach**

| **Q (1-3)** |
|---|
| "I usually do electrostimulation right after school. I don't waste much time because I can read a book or study while the muscles do it by themselves." (P18)<br><br>"Yes, when you just do them at home, with two balls, mainly for the hands, which I do. That's actually mostly what I do at home. And I do it in front of the TV as well." (P15)<br><br>"We came home from [name of a city] yesterday. I drove myself and can drive myself. It went quite well…I can do it while catheterized. Then it doesn't bother anyone. It's much easier for me, and I feel freer. I don't have to wait to find a clean restroom" (P17) |

**Q (4-8)**

"I only do that once a week. I have a supply box, I prepare it, that's more on the side. When I finish everything, a good hour is gone. But that's a day I can use for something else. Because if I sit down every morning to organize the medications, I don't have the time for that, because I still have to go to training and so on. Besides, it's annoying. It's really better to sit down once and get it done." (P29)

"I actually enjoy cooking and usually do it on weekends when I have time. I make large portions so that I can eat them throughout the week." (P12)

"I drink a little less now because there isn't always a toilet nearby. If I have a long car journey, I make sure not to drink too much beforehand." (P23)

"I just have to make sure I have certain things with me. Like the citric acid or an absorbent pad when I notice that... Well, I have a completely spastic bladder, and it can suddenly contract, rendering everything useless. It just tightens up. So, two or three drops. But without a pad, it would be wet underneath and uncomfortable. Things like that... Or maybe I'd pack a urine bag if I go to the cinema or make one myself from Bavaria. If I'm going to a three-hour movie, I do that. But otherwise... It's mostly about planning ahead and dealing with bladder-related issues." (P4)

**Q (10-12)**

"I'm part of a wagon-building group…Ultimately, I see the whole wagon-building process, when it starts again, as an exercise, as training. Because these are all movements I might not make in my normal daily life" (P19)

"Yes, household chores also come into play. That's like therapy too. Vacuuming. And yes, there are also tasks that I don't particularly enjoy, like dusting and dishwashing." (P12)

"The doctor told me, I could do physiotherapy…I have a lot of fun at home. I ride at home or go out with friends. I always do transfers. For me, it's more fulfilling than going to a fitness center..." (P28)

**Q (13-15)**

"Yes, that's right. I'm fortunate to have that support. My wife works, and I'm often alone for lunch. But things are already prepared, and we discuss beforehand what I can eat. That's actually quite helpful." (P3)

"My partner takes care of the entire household when it comes to household chores. She handles that. Well, I cook now too. But she does a lot more... We've decided to have groceries delivered, almost everything. So, we don't have to go out ourselves." (P5)

"Spitex prepares my medications, and I have a pill organizer at home, and they fill it once a week. Then they come and do that. My husband helps me prepare it in the evening and morning, so he does the majority of it. Without him, I'd be in a tough spot, I have to say. He's a big help. " (P9)

Spitex-home service
