## Supplemental_4 for "Approaches to self-management integration and influencing factors in everyday life after spinal cord injury: A qualitative narrative analysis"

**Quotes representing the three different groups of factors influencing SM integration**

**Table 1. Mind and body dynamics**

| **Q (1)** |
|---|
| "Because medications make me tired. Depending on what I do during the day, they make me tired to varying degrees. And I'm still figuring out how much energy I need for various things… if I have absolutely no energy left, then I need to sleep. It could even be at 6 PM…I now know, for example, that when I go to the office, I need to stop early, go home, so that I can still do something at home in the evening. Because I need energy for that, so that I'm not completely drained by then." (P7) |
| **Q (2-3)** |
| "I would have done the same exercises there [at home], but I didn't manage, as I mentioned earlier, with the other things that kept me busy, everything that was new and different... it takes so much...Brain power. That's also the reason why I couldn't concentrate anymore or why I forgot everything." (P3)<br><br>"Yes, I would say that what I find most challenging is this small mental, psychological aspect. Because somehow, you need time to navigate between what I can do, what is possible, what I want, and what I don't want." (P13) |

**Table 2. Environmental and informational dynamics**

| **Q (1-8)** |
|---|
| "It's a bit of everything. I live alone and do everything myself. So, whatever needs to be done at home, I have to do. So, it really affects me throughout." (P7)<br><br>"But the best part was that I wasn't alone during the first few days of getting used to being back home. I believe it would have been much more challenging for my mental state if I had gone from being in the protected environment of the [rehabilitation center] and being part of a large, cool group to suddenly living alone. It would have been like going from 100 to 0 overnight". (P19)<br><br>"They also check if the bowel is empty, so it takes two days until it comes again. If there's something in there, it's more difficult. I can clean, but I can't see what I'm cleaning. That's annoying… The problem is always that I need help. That's my problem. They come at 6…It's always a bit stressful." (P2)<br><br>"So, you have to be a bit self-reliant. Spitex is also an option, of course. You can call them, and they will organize and provide everything. You don't have to do much…Because with Spitex, they even come to wash the person themselves, administer medications, and so on. I only needed household help, and that too, at most twice a week. That's what we agreed upon." (P12)<br><br>"Can we stop home care service? That's the main hurdle. In the evening, with home care service, you're less free than in the morning. That's quite significant. But in the evening, you always have to be at home on time. You don't know when home care service will come. They |

| |
|---|
| come between 7 and 10 in the evening. That's uncomfortable." (P16)<br><br>"The sessions [therapy] are in the middle of the afternoon, which is a bit annoying because you can't do anything else. You can't say, "I'll go out in the early afternoon, and at 3 pm, I'll be there." Then, after 3 pm, you have almost two hours at home, and by 5 pm, it's dark…the afternoon is dead… It's the same with the nurse. She comes at 3 pm. So, it's a bit annoying." (P32)<br><br>"Then the renovation came up. I was lucky about that. I've been living in the same house for over 40 years…It's 150 m2 on one level. It's like an apartment where everything is on the same level…The doors are flat at the bottom. We didn't need to modify the toilet. We had to modify the shower. There was a bathtub there. Now I have a 120 cm wide shower. (P8)<br><br>"They also looked at the shower, how I get in here. They wanted to remodel the kitchen…I set up a table and made sure I can maneuver around it. I prepare and cook here. I have a hot plate there. I have a Thermomix, which is great, of course. Genius. Then I can do my things here on the table. I don't have to... If I have to wash something by hand, I wash it. It's always a bit tedious to empty it, but it works." (P2) |
| **Q (9-12)** |
| "At some point, I realized that, for example, I couldn't sweat in the gym. I could strain and do whatever I wanted, but I wouldn't sweat…I wasn't prepared on how to handle myself in the summer. You have to make sure there's shade. If there's something I need to do more it's sun rest… the preparation for the sun…I never really got fully prepared for it." (P16)<br><br>"One thing that comes to mind is sexuality. It's all about sexuality… But sexuality always involves two people. It can't be that my wife is a sparring partner for my sexuality…It's not that simple. I believe that apart from the question of whether there is sexuality, whether there is an erection, it's always preceded by the assumption that a partner is completely available. That's not the case. That's one of the aspects I have to mention. In urology, they may be more open…But it's actually a taboo…That sexuality is, at least, a partnership where someone doesn't just come into a seminar…this workshop came relatively early for me. It's a must. That was okay. But my wife didn't attend because she finds it repulsive." (P17)<br><br>"What I find helpful is that a group formed during rehab. We still have contact. Very different physical conditions. But that's something I appreciate extremely…I found the peer exchange moderately helpful. At first, I had someone who was a tetraplegic. I canceled it…..That was a bit difficult. It's not bad to learn from the peers. That's not bad. I find it very useful. But the individual peer sessions didn't bring me much. The one-to-ones. With those I met, it was different. From the physical conditions, everyone is different. Now you've started working again. I find that useful. I don't know if that can be organized. Or if it's something that runs self-organized. I find it useful. You meet with people you get along well with." (P30)<br><br>"Yes, we have a WhatsApp group where someone writes to us. Then I share with you a video that you can experience…That's something that benefits me. The real possible possibilities. If I had had the accident 20 years ago, I wouldn't have survived because I can't see it. And secondly, you can be in contact with the phone, with WhatsApp, with other people" (P20) |

**Table 3. Society and perception dynamics**

| **Q (1-2)** |
|---|
| "I'm about to start working a bit again…when I go to the toilet, it won't be done in 5 minutes. It takes about fifteen minutes or so. So no one should be surprised if I disappear for 20 minutes. That's just how it is." (P29)<br><br>"Yes. Work is basically not a problem. I've told the boss, if things get too much for me, I might just retreat to the car, lean back and relax for a bit. They also said it wouldn't be a problem…I feel like you could take a break, if needed. Because of the flexible working hours and the guided working hours, I can manage very well… If something's wrong, I have to say quickly, then I'll pull back or go home earlier. I want to relax." (P25) |
| **Q (3-6)** |
| "When someone asks if I had an accident, I say, well, not exactly, no. I actually had a herniated disc, had surgery, woke up with paralysis, and had to go to rehabilitation to learn to walk again. And then they're like, oh, now you're standing, now you're walking (when I do the exercises and movements)... now you're fixed, you should be able to walk, so why do you still need a wheelchair? And deep down, I feel like I shouldn't care what others think, but at the same time, it's not always easy to ignore their opinions…I feel like people might think I'm faking or taking advantage of the help." (P13)<br><br>"People are extremely surprised when you come on a bike. I get off or on almost elegantly. Then I take the crutches. Then I wobble. From the bike to the store. You don’t see that often. I think it needs more awareness in society. There is not just black and white, but also a lot in between…I don’t know if it needs more awareness. The family doctor said it’s a rarity… But that being in a wheelchair doesn’t mean always being in a wheelchair, that’s noticeable to me. If someone is in a wheelchair, their biggest problem usually isn’t that they can’t walk, but that bladder and bowel management is a huge issue. People don’t always get that. I sometimes ask, if you can walk, why are you in a wheelchair? The wheelchair has such a stigma that many people can’t imagine being in it voluntarily…The way you walk isn’t any healthier. But it’s so stigmatized, that’s true." (P30)<br><br>"Because there are many things from the outside, they don't see that. They don't see that you need a catheter. I remember here, colleagues visited me. And we sat together, drank cola and so on. And then I had to go to the toilet. Then I said, it takes a little longer. And afterward, they asked why it took so long. Then I showed them the catheter. They almost fled." (P29)<br><br>"When I had incontinence, we made an arrangement. I had to say, maybe not today. I had colleagues who understood. They said, we'll cook at your place." (P21) |
